# The association between multidimensional sleep health and gestational weight gain: nuMoM2b Sleep Duration and Continuity Study

**DOI:** 10.1101/2023.02.21.23285931

**Authors:** Marquis S. Hawkins, Darya Y. Pokutnaya, Lisa M. Bodnar, Michele D. Levine, Daniel J. Buysse, Esa M. Davis, Meredith L. Wallace, Phyllis C. Zee, William A. Grobman, Kathryn J. Reid, Francesca L. Facco

## Abstract

**Background:** Although poor sleep health is associated with weight gain and obesity in the non-pregnant population, research on the impact of sleep health on weight change among pregnant people using a multidimensional sleep-health framework is needed. This study examined associations among mid-pregnancy sleep health indicators, multidimensional sleep health, and gestational weight gain (GWG).

**Methods:** We conducted a secondary data analysis of the Nulliparous Pregnancy Outcome Study: Monitoring Mothers-to-be Sleep Duration and Continuity Study (n=745). Indicators of individual sleep domains (i.e., regularity, nap duration, timing, efficiency, and duration) were assessed via actigraphy between 16 and 21 weeks of gestation. We defined “healthy” sleep in each domain based on empirical thresholds. Multidimensional sleep health was based on sleep profiles derived from latent class analysis. Total GWG, the difference between self-reported pre-pregnancy weight and the last measured weight before delivery, was converted to z-scores using gestational age- and BMI-specific charts. GWG was defined as low (<−1 SD), moderate (−1 or +1 SD), and high (>+1 SD).

**Results:** Nearly 50% of the participants had a healthy sleep profile (i.e., healthy sleep in most domains), whereas others had a sleep profile defined as having varying degrees of poor health in each domain. While indicators of individual sleep domains were not associated with GWG, multidimensional sleep health was related to low and high GWG. Participants with a sleep profile characterized as having low efficiency, late timing, and long sleep duration (vs. healthy sleep profile) had a higher risk (RR 1.7; 95% CI 1.0, 3.1) of low GWG a lower risk of high GWG (RR 0.5 95% CI 0.2, 1.1) (vs. moderate GWG).

**Conclusions:** Multidimensional sleep health was more strongly associated with GWG than individual sleep domains. Future research should determine whether sleep health is a valuable intervention target for optimizing GWG.

**Synopsis:** *Study question:*

- What is the association between mid-pregnancy multidimensional sleep health and gestational weight gain?

*What’s already known?:*

- Sleep is associated with weight and weight gain outside of pregnancy

*What does this study add?:*

- We identified patterns of sleep behaviors associated with an increased risk of low gestational weight gain

## Background

Low or high gestational weight gain (GWG), defined by the Institute of Medicine (IOM) recommendations,^1^ is associated with adverse maternal outcomes (e.g., cesarean section, gestational diabetes, hypertensive disorders of pregnancy, postpartum weight retention)^2^ and child outcomes (e.g., insulin resistance, small-for-gestational-age, large-for-gestational-age, shoulder dystocia, childhood adiposity).^2–6^ The US Preventive Service Task Force recommends that clinicians offer all pregnant people behavioral counseling interventions to promote healthy GWG. Most behavioral counseling interventions for pregnant people address diet and physical activity. However, these standard interventions are only modestly effective. For example, a systematic review and meta-analysis by the US Preventive Service Task Force showed that diet and physical activity interventions (vs. control) only resulted in one kg lower total GWG and were not associated with adherence to the IOM recommendation (neither gaining too much nor too little weight).^7^ Thus, there is a need to identify additional targets for lifestyle interventions that can promote healthy GWG.

Sleep health is a promising intervention target to manage GWG. Poor sleep health may lead to weight gain through hormonal (e.g., dysregulation of leptin and ghrelin)^8–11^ and behavioral (e.g., increased food consumption, decreased physical activity) pathways.^12–17^ In non-pregnant populations, poor sleep health indicators such as short or long sleep duration, early or late sleep timing, low sleep efficiency (a measure of continuous vs. interrupted sleep), and low sleep regularity across days are independently associated with obesity and weight gain.^18^ Among the few studies examining associations between sleep and GWG, most,^19–21^ but not all, ^22^ studies have found short sleep duration and perceived sleep deprivation to be associated with increased risk of low GWG (i.e., less than NAM guidelines).

The research examining associations between sleep and GWG is limited for several reasons. First, prior research primarily focuses on associations of two specific measures of sleep -- duration and self-reported quality, but not other sleep domains (e.g., sleep timing and regularity) associated with obesity in non-pregnant populations. Second, sleep health is best characterized by multiple co-occurring domains that together are more strongly associated with health outcomes than individually.^23, 24^ However, prior studies in pregnancy examined the associations of sleep health indicators and GWG individually in separate models. Lastly, most studies relied on self-reported measures of sleep, which can overestimate (e.g., sleep duration) or underestimate (e.g., wake after sleep onset) sleep in some domains.^25^ Studies with objective measures of sleep using actigraphy can more accurately measure sleep health, which would further our understanding of the association between sleep health and GWG.

In light of the current research gaps, this study aimed to 1) characterize mid-pregnancy multidimensional sleep and 2) examine associations between indicators of individual sleep domains, multidimensional sleep, and GWG. We hypothesized that multidimensional sleep health indicator is more strongly associated with GWG than individual sleep health indicators.

## Methods

### Study Design & Population

The nuMoM2b (Nulliparous Pregnancy Outcome Study: Monitoring Mothers-to-be) Pregnancy and Sleep Duration and Continuity Study is ancillary to the parent nuMoM2b study.^26^ The nuMoM2b was a prospective cohort study to identify maternal characteristics and environmental factors predictive of adverse pregnancy outcomes in nulliparous women.^26^ The study enrolled 10,038 nulliparous women (no prior delivery >20 weeks’ gestation) with a viable singleton pregnancy at the time of screening (6 to 13 weeks of gestation; Visit 1) from eight clinical sites across the US. Three additional assessments occurred at 16 to 21 weeks of gestation (Visit 2), 22 to 31 weeks of gestation (Visit 3), and delivery. In the sleep duration and continuity ancillary study, a subset of participants (n=901) was asked to complete a 7-day sleep assessment using a wrist actigraphy monitor and sleep diary at the visit 2 assessment. This ancillary study aimed to determine the association between objectively measured sleep and pregnancy-related cardio-metabolic morbidity.

The sleep duration and continuity study excluded nuMoM2b study participants younger than 18 years of age because sleep characteristics differ between adults and children. Participants were also excluded if they had pre-existing chronic hypertension (n=8) or diabetes (n=26) since the study aimed to diagnose the new onset of these conditions in pregnancy. For this analysis, we further excluded participants who had invalid sleep data (n=119) (described further below) and missing GWG values (n=37). The final analytic sample included 747 participants. Each study site obtained IRB approval, and each participant provided informed consent before assessments.

### Sleep Assessment

Reid et al. described the quality control procedures and actigraphy scoring for the sleep duration and continuity sub-study.^27^ Briefly, participants were asked to wear the actigraphy monitor on their non-dominant wrist for 7 consecutive days. Participants were instructed to press the monitor’s event marker button at their primary bedtime (i.e., when turning off the light or trying to fall asleep), primary wake time, and when taking naps or falling asleep for more than 5 minutes. Also, participants completed a log to help interpret the actigraphy data.

A single trained technician scored all actigraphy data at a central reading center. The technician manually set the rest intervals using a standardized formula based on the actigraphy event markers and sleep logs. The technician used activity or light levels recorded by the monitor if an event marker and log were unavailable. A second trained technician scored the actigraphy data if there were small (i.e., <30 minutes) differences in the rest intervals between the event markers and sleep logs or activity/light levels. The reading center’s director adjudicated disagreements between the technicians. The reading center director decided on rest interval placement for large (>30 minutes) differences in the rest intervals between the event markers and sleep logs or activity/light levels. An actigraphy recording was considered valid if, during the 7 days of the study, there were at least 5 primary sleep periods recorded (out of a possible 7), there were less than 4 hours of off-wrist time during the 24-hour periods containing the primary sleep periods, and there was no off-wrist time during the sleep period. If a participant’s study data did not meet these criteria, they were asked to wear the watch again for an additional week as long as the recording could be completed by 23 weeks of gestation. The actigraphy data were configured using Actiware Sleep V5.59 (Phillips-Respironics, Mini Mitter, Bend, OR) to record epochs with a 30-second duration and the presence of red, green, and blue light. The activity threshold was set at medium (i.e., 40 activity counts).

We selected indicators of sleep domains that map to Buysse’s sleep health framework^28^ and that can be derived from actigraphy.^29^ We used National Sleep Health Foundation recommendations or thresholds used in prior studies to define “healthy” sleep in each domain.^30–32^ *Sleep timing* was based on the sleep midpoint, the halfway point between the participants’ sleep onset and final wake-up time to start the day (i.e., sleep offset). Healthy sleep health was defined as a sleep midpoint between 2:00 am and 4:00 am. Early sleep timing was defined as a sleep midpoint < 2:00 am, and late sleep timing was defined as a sleep midpoint after 4:00 am. *Sleep regularity* was defined as the standard deviation difference in sleep midpoint during the 7-day assessment period. Healthy sleep regularity was defined as less than a one-hour standard deviation. *Sleep duration* was estimated by taking the weighted average of weekday/workday and weekend/non-workday total sleep duration during the primary (usually night-time) sleep period. We defined healthy sleep in this domain as 7 to 9 h of sleep per night.^33^ Short sleep duration was defined as less than 7 h of sleep per night. Long sleep duration was defined as greater than 9 h of sleep each night. *Sleep efficiency* was defined as the proportion of actual sleep time within the period from sleep onset to sleep offset to start the day; high sleep efficiency indicates less wakefulness during the night. We defined healthy sleep efficiency as spending at least 85% of the total sleep interval asleep. *Napping* was defined as falling asleep for at least 15 minutes during the day. We calculated the average frequency and duration of naps during the assessment period. We defined a healthy nap duration as <100 minutes per day. We set nap frequency and duration to zero for participants who did not nap. *Multidimensional sleep health* was calculated using two methods described in the statistical analysis section.

### Covariates and Descriptive Factors

Participants self-reported their demographic characteristics, including age, the highest level of educational attainment (<college, some college and beyond), and race and ethnicity at visit 1. The Edinburgh Postnatal Depression Scale (EPDS) was used to measure depressive symptoms at Visit 1. The EPDS has good sensitivity (86%) and specificity (78%) for identifying perinatal depression risk.^34^ Participants self-reported smoking status three months before pregnancy. Participants self-reported their frequency and duration of exercise (e.g., running, aerobics, gardening, walking for exercise) in the past 4 weeks. Participants reported their distance travel for running, jogging, walking, swimming, or cycling activities. We calculated the total duration per week for all reported activities. Diet quality was assessed with a food frequency questionnaire. We calculated the Healthy Eating Index values to estimate the overall dietary quality.^35^

### Primary Outcome: Gestational Weight Gain

Total GWG was defined as the difference between self-reported pre-pregnancy weight and the chart-abstracted last measured weight before delivery. Total weight gain was converted to z-scores using gestational age- and BMI-specific charts.^36^ Converting absolute total weight gain to gestational age-standardized z-scores produces a measure uncorrelated with gestational age at delivery, removing bias based on gestational age at birth. Because low and high GWG is associated with adverse maternal and child outcomes,^37^ we used Z-scores to define low (<−1 SD), moderate (−1 or +1 SD), and high (>+1 SD) total GWG. We eliminated data from participants with extreme GWG values defined as z-scores < −5 or > 5 SD (n=7) because of concern that they represented measurement error.

### Statistical Analysis

For aim 1, we used two methods that have been used in prior research to calculate multidimensional sleep health.^24, 32, 38, 39^ First, we calculated multidimensional sleep health using a simple composite score, an aggregate of “healthy” sleep indicators in each domain. For simplicity, we will refer to the composite score as an estimate of overall sleep health. The score ranges from 0 to 5, with higher scores suggesting better sleep health.

Second, we defined multidimensional sleep health based on sleep patterns/profiles derived from latent class analysis (LCA). LCA assumes a finite number of latent (i.e., unmeasured) classes whose distributions are a combination of measured nominal variables (i.e., sleep health indicators described above). The latent classes represent meaningful subgroups (i.e., sleep profiles) that explain variable clusters. Participants are assigned to the latent class for which they have the highest membership probability. To determine the number of latent classes that fit the data best, we fit a sequence of models, starting with a one-class structure and then adding one class at a time. We used the Bayesian Information Criterion (BIC) and Akaike Information Criterion (AIC) to assist in selecting the final class structure. A better fit is indicated by lower BIC and AIC values. We then used a bootstrap likelihood ratio test to compare the fit of models with fewer classes. We identified four latent classes (i.e., sleep profiles). Supplemental Table 2 provides the AIC and BIC values for each class and the p-value from the bootstrap likelihood ratio test comparing the fit of models with fewer classes. The bootstrap LRT test indicated that a 3-class structure best fit the data (3-class vs. 2-class, p<0.01; 4-class vs. 3-class, p=0.23). We will use the class with the best sleep profile as the reference group for all models.

For aim 2, the primary analysis, we used multinomial logistic regression to examine the association between the indicators of individual sleep health domains and multidimensional sleep health to determine the odds of low or high GWG (moderate GWG was the reference group). We fit multiple models, first unadjusted and then controlling for demographic factors.^40, 41^ We used a directed acyclic graph to identify covariates for multivariable adjustment. First, we identified empirical predictors of gestational weight gain.^40–42^ Then, we selected factors associated with GWG that were also associated with sleep health based on our expertise. Specifically, we adjusted for descriptive covariates, including age, mid-pregnancy depressive symptoms, education, marital status, and smoking status.

We generated descriptive and regression tables using the summary package in R (version 1.5.2). Latent class analyses were performed using the poLCA package in the R statistical environment (R version 4.0.2, RStudio Version 1.3.1073). STATA SE was used to derive relative risk ratios from the multinomial regression models (StataCorp, version 16, College Station, TX).

## Results

A total of 901 people in the nuMoM2b parent study were enrolled in the Sleep Duration and Continuity Substudy. Of the 901 people who participated in the sub-study, 782 (87%) had valid actigraphy sleep data. Among the 119 people with invalid sleep data, the two most frequent reasons were participant noncompliance (60%, watch not worn at least 20 hours/day for at least 5 days) and watch failure (40%). Watch failure was primarily due to one of the following reasons: faulty off-wrist detection, a corrupted database, or constant low-level activity counts.^27^ A total of 759 individuals had sleep diary data. After additionally removing participants with missing or extreme GWG values (n=7), the final analytic sample included 745 participants.

Table 1 provides the final analytical sample’s demographic characteristics. The sample had a median age of 27 years, was predominantly white, college educated, and currently employed. Among individuals currently employed, approximately 20% work an afternoon, night, or irregular shift. The median GWG was 15 kg (IQR 12, 20) (Table 1). The median GWG among individuals in the low GWG group was 8 kg, individuals in the moderate GWG group were 15 kg (IQR 13, 18), and individuals in the high GWG group were 25 kg (IQR 23, 28).

**Table 1.** Sample descriptive characteristics

| Table 1 - Sample descriptive characteristics |  |
| --- | --- |
| Characteristic | N = 745<sup>1</sup> |
| Age, yrs | 27 (23, 31) |
| Gestational Age | 39.00 (38.00, 40.00) |
| Race/Ethnicity |  |
| Non-Hispanic White | 478 (64%) |
| Non-Hispanic Black | 88 (12%) |
| Hispanic | 112 (15%) |
| All Other Race/Ethnicities | 67 (9.0%) |
| Education |  |
| <College | 278 (37%) |
| Some College or Beyond | 467 (63%) |
| Work schedule |  |
| Not working | 145 (21%) |
| Day shift | 416 (59%) |
| Afternoon/Night/Irregular shift | 142 (20%) |
| Income Level |  |
| High | 441 (59%) |
| Middle | 85 (11%) |
| Low | 85 (11%) |
| Unknown | 134 (18%) |
| Edinburgh Postnatal Depression Score | 5.0 (2.0, 8.0) |
| Pre-pregnancy BMI | 23.9 (21.3, 27.6) |
| MVPA, min/day | 120 (0, 360) |
| Health Eating Index | 65 (54, 73) |
| Pre-pregnancy Smokers | 110 (15%) |
| Gestational weight gain, kg | 15 (12, 20) |
| Characteristic | N = 745<sup>1</sup> |
| <sup>1</sup>Median (IQR); n (%) |  |

### Sleep health characteristics

Overall, participants had healthy sleep in a median of three sleep domains. Most participants had healthy sleep duration, with a median of 8.2 hours/night (IQR 7.7, 8.7). Only 24% (n = 177) of participants reported napping more than 100 minutes/per day. A lower proportion of people had healthy sleep efficiency and timing than other sleep domains. Specifically, 48% (n = 386) of participants had poor sleep efficiency, and 38% (n = 438) had late sleep timing (Table 2).

**Table 2.** Sleep characteristics overall and by sleep profile

| Characteristic | Sleep Profiles |  |  |  |  |
| --- | --- | --- | --- | --- | --- |
|  | Overall, N = 745<sup>1</sup> | Profile 1, N = 362<sup>1</sup> | Profile 2, N = 204<sup>1</sup> | Profile 3, N = 119<sup>1</sup> | Profile 4, N = 60<sup>1</sup> |
| Sleep midpoint regularity | 49 (35, 68) | 44 (33, 58) | 59 (41, 84) | 49 (32, 67) | 81 (51, 100) |
| Sleep midpoint, SD <60 min | 491 (66%) | 285 (79%) | 105 (51%) | 81 (68%) | 20 (33%) |
| Nap duration, min | 55 (0, 98) | 44 (0, 78) | 68 (0, 112) | 48 (0, 86) | 104 (80, 154) |
| Nap Duration, <100 min/day | 568 (76%) | 308 (85%) | 140 (69%) | 93 (78%) | 27 (45%) |
| Sleep Timing | 3:38am (2:56am, 4:38am) | 3:07am (2:41am, 3:32am) | 4:46am (4:16am, 5:51am) | 4:12am (3:16am, 5:14am) | 4:01am (3:11am, 6:04am) |
| Sleep Timing |  |  |  |  |  |
| <2am | 22 (3.0%) | 0 (0%) | 20 (9.8%) | 2 (1.7%) | 0 (0%) |
| 2-4am | 435 (58%) | 359 (99%) | 0 (0%) | 48 (40%) | 28 (47%) |
| >4am | 288 (39%) | 3 (0.8%) | 184 (90%) | 69 (58%) | 32 (53%) |
| Sleep Efficiency, % | 85.3 (81.7, 88.5) | 86.4 (83.3, 89.2) | 84.6 (80.6, 87.8) | 83.7 (80.2, 87.4) | 84.0 (79.7, 87.1) |
| Sleep efficiency, >85% | 386 (52%) | 217 (60%) | 98 (48%) | 46 (39%) | 25 (42%) |
| Sleep Duration, h/night | 8.16 (7.67, 8.69) | 8.06 (7.71, 8.38) | 8.09 (7.76, 8.55) | 9.39 (9.15, 9.77) | 6.71 (6.33, 6.89) |
| Sleep Duration, min/day |  |  |  |  |  |

Table 2- Sleep characteristics overall and by sleep profile
| Characteristic | Sleep Profiles |  |  |  |  |
| --- | --- | --- | --- | --- | --- |
|  | Overall, N = 745<sup>1</sup> | Profile 1, N = 362<sup>1</sup> | Profile 2, N = 204<sup>1</sup> | Profile 3, N = 119<sup>1</sup> | Profile 4, N = 60<sup>1</sup> |
| Short Sleep | 60 (8.1%) | 0 (0%) | 0 (0%) | 0 (0%) | 60 (100%) |
| Sufficient Sleep | 566 (76%) | 362 (100%) | 204 (100%) | 0 (0%) | 0 (0%) |
| Long Sleep | 119 (16%) | 0 (0%) | 0 (0%) | 119 (100%) | 0 (0%) |
| Global Sleep Health | 3.0 (2.0, 4.0) | 4.0 (3.0, 4.0) | 2.0 (1.0, 3.0) | 2.0 (1.0, 2.0) | 2.0 (1.0, 2.0) |

<sup>1</sup>Median (IQR); n (%)
Profile 1: “Healthy Sleep” – best sleep profile
Profile 2: latest sleep timing
Profile 3: late sleep timing, low sleep efficiency, and long sleep duration
Profile 4: shortest nocturnal sleep duration and longest daytime nap duration

We identified four latent classes that represent distinct sleep profiles. Table 2 depicts the sleep characteristics of each profile. Profile 1 (n=362, 48.6%) had a higher probability of good sleep in each domain compared to all other classes. For simplicity, we will refer to profile 1 as the “healthy sleep” profile. Participants in the healthy sleep profile had healthy sleep in a median of four of five sleep domains, whereas all other profiles had healthy sleep in a median of two sleep domains. Profile 2 (n=204, 27.4%) was best characterized as having the latest sleep timing than the other profiles. Profile 3 (n=119, 16.0%) was best characterized as having late sleep timing, low sleep efficiency, and long sleep duration. Profile 4 (n-60, 8.1%) was best characterized by their short nocturnal sleep duration and long daytime nap duration.

Participants in the healthy sleep profile (vs. all other profiles) were older, reported fewer depressive symptoms, were more physically active and had higher diet quality (Table 3). The distribution of sleep profiles differed by race, education, income level, and smoking status. Specifically, a larger proportion of non-Hispanic White participants had a healthy sleep profile compared to all other racial groups. Likewise, a larger proportion of participants who reported some college or beyond had a high income or worked a day shift had a healthy sleep profile. (Table 3).

**Table 3.** Descriptive characteristics by sleep profile

| Characteristic | Overall, N =<br>745<sup>1</sup> | Profile 1, N =<br>362<sup>1</sup> | Profile 2, N =<br>204<sup>1</sup> | Profile 3, N =<br>119<sup>1</sup> | Profile 4, N =<br>60<sup>1</sup> |
| --- | --- | --- | --- | --- | --- |
| Age, yrs. | 27 (23, 31) | 29 (26, 33) | 25 (21, 30) | 24 (21, 29) | 28 (23, 31) |
| Gestational Age | 39.00 (38.00,<br>40.00) | 39.00 (38.00,<br>40.00) | 39.00 (38.00,<br>40.00) | 39.00 (39.00,<br>40.00) | 39.50 (38.75,<br>40.00) |
| Race/Ethnicity |  |  |  |  |  |
| Non-Hispanic White | 478 (100%) | 275 (58%) | 99 (21%) | 74 (15%) | 30 (6.3%) |
| Non-Hispanic Black | 88 (100%) | 26 (30%) | 35 (40%) | 16 (18%) | 11 (12%) |
| Hispanic | 112 (100%) | 35 (31%) | 49 (44%) | 20 (18%) | 8 (7.1%) |
| All Other Race/Ethnicities | 67 (100%) | 26 (39%) | 21 (31%) | 9 (13%) | 11 (16%) |
| Education |  |  |  |  |  |
| <College | 278 (100%) | 77 (28%) | 102 (37%) | 74 (27%) | 25 (9.0%) |
| Some College or Beyond | 467 (100%) | 285 (61%) | 102 (22%) | 45 (9.6%) | 35 (7.5%) |
| Work schedule |  |  |  |  |  |
| Not working | 145 (100%) | 36 (25%) | 56 (39%) | 41 (28%) | 12 (8.3%) |
| Day shift | 416 (100%) | 265 (64%) | 81 (19%) | 38 (9.1%) | 32 (7.7%) |
| Afternoon/Night/Irregular<br>shift | 142 (100%) | 43 (30%) | 56 (39%) | 31 (22%) | 12 (8.5%) |
| Income Level |  |  |  |  |  |
| High | 441 (100%) | 276 (63%) | 90 (20%) | 43 (9.8%) | 32 (7.3%) |

Table 3 - Descriptive characteristics by sleep profile
| Characteristic | Overall, N = 745<sup>1</sup> | Profile 1, N = 362<sup>1</sup> | Profile 2, N = 204<sup>1</sup> | Profile 3, N = 119<sup>1</sup> | Profile 4, N = 60<sup>1</sup> |
| --- | --- | --- | --- | --- | --- |
| Middle | 85 (100%) | 30 (35%) | 25 (29%) | 22 (26%) | 8 (9.4%) |
| Low | 85 (100%) | 24 (28%) | 31 (36%) | 25 (29%) | 5 (5.9%) |
| Unknown | 134 (100%) | 32 (24%) | 58 (43%) | 29 (22%) | 15 (11%) |
| Edinburgh Postnatal Depression Score | 5.0 (2.0, 8.0) | 4.0 (2.0, 7.0) | 5.0 (3.0, 8.2) | 5.0 (3.0, 9.0) | 5.0 (2.8, 9.0) |
| Pre-pregnancy BMI | 23.9 (21.3, 27.6) | 23.5 (21.2, 27.0) | 24.4 (21.7, 27.8) | 24.9 (21.7, 29.4) | 24.0 (21.7, 31.6) |
| MVPA, min/day | 120 (0, 360) | 190 (60, 479) | 90 (0, 236) | 60 (0, 210) | 78 (0, 360) |
| Health Eating Index | 65 (54, 73) | 69 (59, 75) | 62 (51, 69) | 58 (48, 66) | 59 (50, 69) |
| Pre-pregnancy Smokers | 110 (100%) | 32 (29%) | 38 (35%) | 29 (26%) | 11 (10%) |

<sup>1</sup>Median (IQR); n (%)
<sup>2</sup>Kruskal-Wallis rank sum test; Pearson's Chi-squared test
Profile 1: “Healthy Sleep” – best sleep profile
Profile 2: latest sleep timing
Profile 3: late sleep timing, low sleep efficiency, and long sleep duration
Profile 4: shortest nocturnal sleep duration and longest daytime nap duration

### Sleep Health and Gestational Weight Gain

None of the individual indicators of domain-specific sleep health were associated with GWG in adjusted models (Table 4). Sleep profiles were associated with GWG such that participants with a sleep profile characterized as having low efficiency, late timing, and long sleep duration (vs. healthy sleep profile) had a higher risk (RR 1.7; 95% CI 0.9, 3.1) of low GWG a lower risk of high GWG (RR 0.5 95% CI 0.2, 1.1) (vs. moderate GWG) in models adjusting for education, depressive symptoms, marital status, and smoking status (Table 5). Global sleep health was inversely associated with low GWG risk. Specifically, each additional indicator of good sleep health was associated with a 14% lower risk of having low GWG in adjusted models.

**Table 4.**
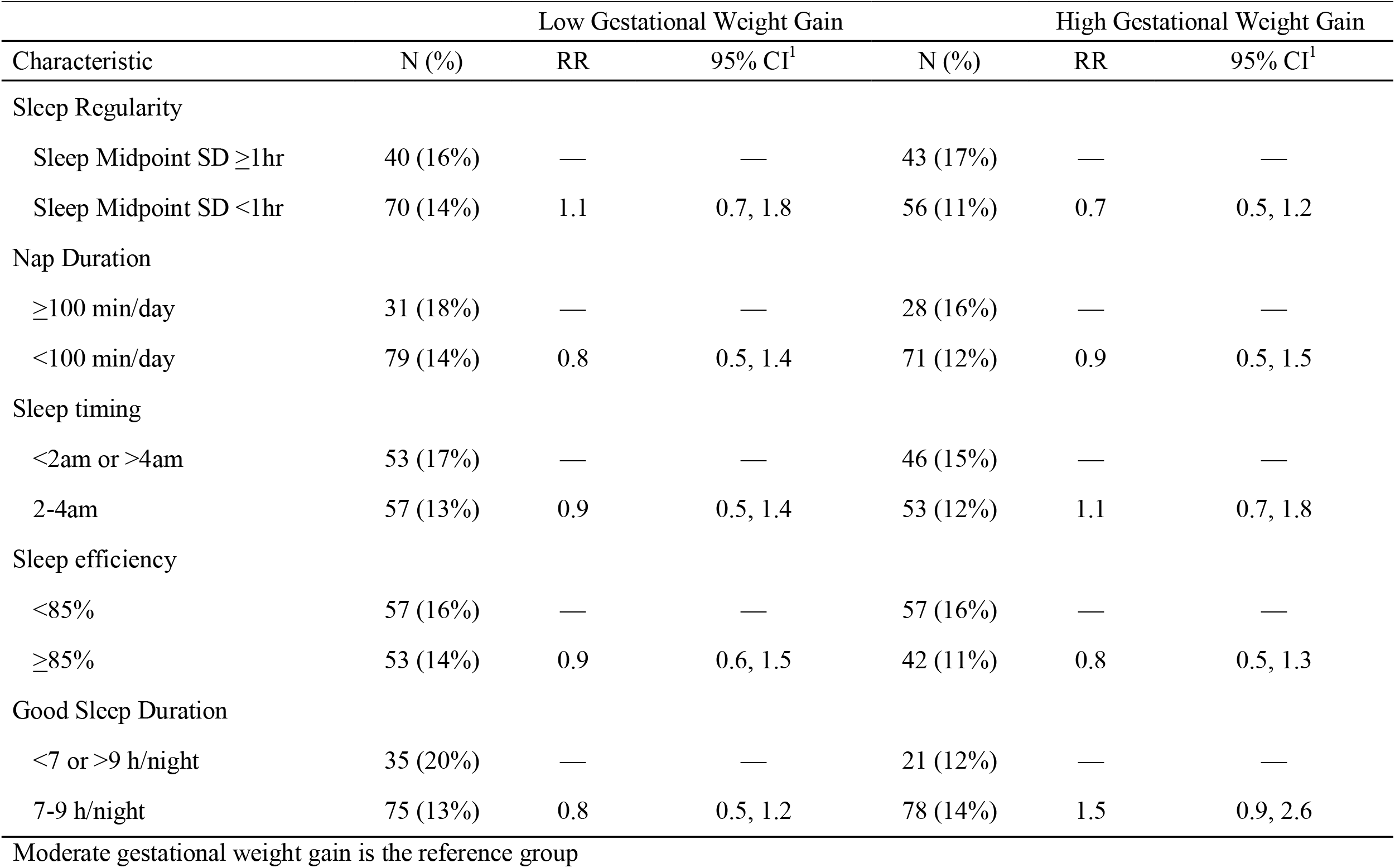
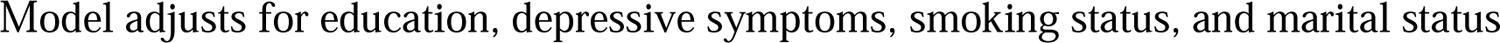
The associations of individual sleep health dimensions and gestational weight gain

| Characteristic | Low Gestational Weight Gain |  |  | High Gestational Weight Gain |  |  |
| --- | --- | --- | --- | --- | --- | --- |
|  | N (%) | RR | 95% CI<sup>1</sup> | N (%) | RR | 95% CI<sup>1</sup> |
| Sleep Regularity |  |  |  |  |  |  |
| Sleep Midpoint SD ≥1hr | 40 (16%) | — | — | 43 (17%) | — | — |
| Sleep Midpoint SD <1hr | 70 (14%) | 1.1 | 0.7, 1.8 | 56 (11%) | 0.7 | 0.5, 1.2 |
| Nap Duration |  |  |  |  |  |  |
| ≥100 min/day | 31 (18%) | — | — | 28 (16%) | — | — |
| <100 min/day | 79 (14%) | 0.8 | 0.5, 1.4 | 71 (12%) | 0.9 | 0.5, 1.5 |
| Sleep timing |  |  |  |  |  |  |
| <2am or >4am | 53 (17%) | — | — | 46 (15%) | — | — |
| 2-4am | 57 (13%) | 0.9 | 0.5, 1.4 | 53 (12%) | 1.1 | 0.7, 1.8 |
| Sleep efficiency |  |  |  |  |  |  |
| <85% | 57 (16%) | — | — | 57 (16%) | — | — |
| ≥85% | 53 (14%) | 0.9 | 0.6, 1.5 | 42 (11%) | 0.8 | 0.5, 1.3 |
| Good Sleep Duration |  |  |  |  |  |  |
| <7 or >9 h/night | 35 (20%) | — | — | 21 (12%) | — | — |
| 7-9 h/night | 75 (13%) | 0.8 | 0.5, 1.2 | 78 (14%) | 1.5 | 0.9, 2.6 |

Moderate gestational weight gain is the reference group
Model adjusts for education, depressive symptoms, smoking status, and marital status

**Table 5.** The associations of multidimensional sleep health dimensions and gestational weight gain

| Characteristic | Low Gestational Weight Gain |  |  | High Gestational Weight Gain |  |  |
| --- | --- | --- | --- | --- | --- | --- |
|  | N (%) | RR | 95% CI<sup>1</sup> | N (%) | RR | 95% CI<sup>1</sup> |
| Global Sleep Health | 110 (15%) | 0.9 | 0.7, 1.0 | 99 (13%) | 1.0 | 0.8, 1.2 |
| Sleep Phenotypes |  |  |  |  |  |  |
| Profile 1 | 42 (12%) | — | — | 44 (12%) | — | — |
| Profile 2 | 33 (16%) | 1.4 | 0.8, 2.3 | 34 (17%) | 1.1 | 0.7, 1.9 |
| Profile 3 | 25 (21%) | 1.7 | 0.9, 3.1 | 10 (8.4%) | 0.5 | 0.2, 1.1 |
| Profile 4 | 10 (17%) | 1.4 | 0.6, 3.1 | 11 (18%) | 1.3 | 0.6, 2.7 |

Moderate gestational weight gain is the reference group
Model adjusts for education, depressive symptoms, smoking status, marital status
Profile 1: “Healthy Sleep” profile, best across each domain
Profile 2: latest sleep timing
Profile 3: late sleep timing, low sleep efficiency, and long sleep duration
Profile 4: shortest nocturnal sleep duration and longest daytime nap duration

## Comment

In this observational cohort of nulliparous pregnant people, we characterized multidimensional sleep health (i.e., global sleep health and sleep profiles) and examined the associations of indicators of individual sleep domains and multidimensional sleep health with GWG. We identified four sleep profiles in the sample population. Approximately 50% of the participants were characterized as having a healthy sleep profile (i.e., healthy sleep health in most assessed domains), with the remainder of participants experiencing varying degrees of disturbances in sleep regularity, nap duration, timing, efficiency, and duration. Consistent with our hypothesis, multidimensional sleep health was more strongly associated with GWG than the indicators of individual sleep domains. Specifically, we found that participants whose sleep profiles were best characterized as having the latest sleep timing, lowest sleep efficiency, and longest sleep duration had a nearly 70% higher risk of low GWG and 50% lower risk of high GWG than participants with a healthy sleep profile. Likewise, each additional healthy sleep indicator was associated with a 14% lower risk of low GWG. In contrast, we did not observe associations between the indicators of individual sleep domains and low GWG risk.

To our knowledge, this study is the first to characterize multidimensional sleep health in pregnancy. Indeed, prior research largely characterizes individual indicators of sleep duration, continuity (e.g., efficiency, sleep onset latency), and quality across pregnancy, with limited data on sleep regularity, timing, napping behaviors, or sleep profiles.^43^ We found that nearly half of the participants had a healthy sleep health profile, and just over half had varying degrees of “unhealthy” sleep regularity, napping behavior, sleep timing, and sleep efficiency. Overall, participants’ sleep efficiency and timing were relatively worse than their sleep health in other domains. Nearly half of the participants had low sleep efficiency (i.e., <85%), and almost 40% had early or late sleep timing. Also, a large proportion of the sample (44%) had low sleep regularity. These findings could have important implications for intervention design because they highlight sleep behaviors most commonly disturbed in mid-pregnancy. Indeed, multiple behavioral intervention strategies, such as cognitive behavioral therapy for insomnia, brief behavioral therapy for insomnia, and sleep education, address these sleep domains and have been shown to improve sleep quality.^44^ Future studies should examine the impact of such interventions on weight management during pregnancy.

Another important finding was that multidimensional sleep health was more strongly associated with low GWG than the indicators of individual sleep domains. This finding supports our a priori hypothesis that the sleep dimensions relate synergistically and suggests that we should consider the patterns of sleep behaviors for assessing the patient’s risk of low gestational weight gain. Unfortunately, there is limited data examining associations of multidimensional sleep health and weight or weight change in women during or outside pregnancy. Hence, it is difficult to contrast this study’s findings with the existing literature. In the Pittsburgh Hill/Homewood Research on Neighborhoods, Sleep, and Health (PHRESH Zzz) study (n=738, 78% female), the investigators did not find cross-sectional associations between actigraphy-derived (i.e., sleep duration, regularity, timing, and efficiency) and self-reported (i.e., sleep satisfaction) indicators of sleep health domains and BMI individually, or as a composite score.^45^ The Study of Women Across the Nation (SWAN) used actigraphy to estimate sleep efficiency, midpoint (timing), length, regularity, and self-reported measures of alertness and satisfaction to construct a composite sleep health score.^32^ They did not find an association between multidimensional sleep health and BMI change after adjusting for age, race, study site, menopausal status, vasomotor symptoms, apnea-hypopnea index, and negative affect. Differences between this study’s findings and the prior literature are likely due to differences in sample characteristics (e.g., age, pregnancy status) and confounder adjustment set.

Our findings that individuals’ sleep domains were not associated with GWG are consistent with the findings of some literature but conflict with others.^19–22, 46, 47^ For example, Abeysena et al. examined associations of self-reported sleep duration in the second and third trimesters and found sleeping <8 h/day (vs. >8 h/day) during the second, third, or both second and third trimesters was associated with higher odds of the IOM defined inadequate GWG (OR 1.60, 95% CI 1.05, 2.46) adjusting for the effect of body mass index and gestational age.^19^ Likewise, Hill et al. found that short sleep duration in late pregnancy was associated with higher odds of inadequate GWG (1.39; 1.03–1.87) and inversely associated with gestational fat gain (−0.36 – 0.16 0.02).^20^ Hill et al. also found that lower sleep quality, assessed with the Pittsburgh Sleep Quality Index, was associated with higher GWG. However, we did not find associations between individual sleep health indicators and GWG. Differences between this study’s findings and prior literature are likely due to sampling variability, sleep assessment timing (assessment in prior studies ranged between 12 to 28 weeks of gestation), assessment methods (e.g., self-report, actigraphy), and outcome assessment (IOM recommendations based on absolute total weight gain vs. gestational age-standardized weight gain z-scores).

There are limitations to consider when interpreting the study findings. As mentioned, the sleep health profiles identified in this study were derived from available sleep measures at visit 2. Sleep characteristics change over pregnancy, and the patterns of change may significantly impact GWG. It is also important to consider that the sleep profiles we identified depend on specific indicators of the sleep domains we assessed. Thus, the number and characteristics of the sleep profiles identified in the current study may differ from what is reported in other samples. Lastly, total GWG was measured using self-reported pre-pregnancy weight, which could lead to under-reporting.

In conclusion, multidimensional sleep health is more strongly associated with low GWG than indicators of individual sleep health domains. Future research should consider sleep health profiles during pregnancy when assessing patients’ risk for low GWG. Future research should identify predictors of good sleep profiles and determine if interventions that promote healthier sleep profiles can promote healthier GWG.

## Data Availability

Data derived from the nuMoM2b study are available in the NICHD DASH repository

## Acknowledgments

None

## References

1. American College of O, Gynecologists. ACOG Committee opinion no. 548: weight gain during pregnancy. Obstet Gynecol. 2013;121(1):210–2. Epub 2012/12/25. doi: http://10.1097/01.AOG.0000425668.87506.4c10.1097/01.aog.0000425668.87506.4c. PubMed PMID: 23262962.

2. LifeCycle Project-Maternal O, Childhood Outcomes Study G, Voerman E, Santos S, Inskip H, Amiano P, Barros H, Charles MA, Chatzi L, Chrousos GP, Corpeleijn E, Crozier S, Doyon M, Eggesbo M, Fantini MP, Farchi S, Forastiere F, Georgiu V, Gori D, Hanke W, Hertz-Picciotto I, Heude B, Hivert MF, Hryhorczuk D, Iniguez C, Karvonen AM, Kupers LK, Lagstrom H, Lawlor DA, Lehmann I, Magnus P, Majewska R, Makela J, Manios Y, Mommers M, Morgen CS, Moschonis G, Nohr EA, Nybo Andersen AM, Oken E, Pac A, Papadopoulou E, Pekkanen J, Pizzi C, Polanska K, Porta D, Richiardi L, Rifas-Shiman SL, Roeleveld N, Ronfani L, Santos AC, Standl M, Stigum H, Stoltenberg C, Thiering E, Thijs C, Torrent M, Trnovec T, van Gelder M, van Rossem L, von Berg A, Vrijheid M, Wijga A, Zvinchuk O, Sorensen TIA, Godfrey K, Jaddoe VWV, Gaillard R. Association of Gestational Weight Gain With Adverse Maternal and Infant Outcomes. JAMA. 2019;321(17):1702–15. Epub 2019/05/08. doi: 10.1001/jama.2019.3820. PubMed PMID: 31063572; PMCID: PMC6506886.

3. Catalano PM, Presley L, Minium J, Hauguel-de Mouzon S. Fetuses of obese mothers develop insulin resistance in utero. Diabetes Care. 2009;32(6):1076–80. doi: 10.2337/dc08-2077. PubMed PMID: 19460915; PMCID: 2681036.

4. Howell KR, Powell TL. Effects of maternal obesity on placental function and fetal development. Reproduction. 2017;153(3):R97–R108. Epub 2016/11/20. doi: 10.1530/REP-16-0495. PubMed PMID: 27864335; PMCID: PMC5432127.

5. Dude AM, Grobman W, Haas D, Mercer BM, Parry S, Silver RM, Wapner R, Wing D, Saade G, Reddy U, Iams J, Kominiarek MA. Gestational Weight Gain and Pregnancy Outcomes among Nulliparous Women. Am J Perinatol. 2021;38(2):182–90. Epub 2019/09/07. doi: 10.1055/s-0039-1696640. PubMed PMID: 31491800; PMCID: PMC7071163.

6. Catalano PM, Shankar K. Obesity and pregnancy: mechanisms of short term and long term adverse consequences for mother and child. BMJ. 2017;356:j1. Epub 2017/02/10. doi: 10.1136/bmj.j1. PubMed PMID: 28179267; PMCID: PMC6888512.

7. Shieh C, Cullen DL, Pike C, Pressler SJ. Intervention strategies for preventing excessive gestational weight gain: systematic review and meta-analysis. Obes Rev. 2018;19(8):1093–109. Epub 2018/05/29. doi: 10.1111/obr.12691. PubMed PMID: 29806187.

8. Rihm JS, Menz MM, Schultz H, Bruder L, Schilbach L, Schmid SM, Peters J. Sleep Deprivation Selectively Upregulates an Amygdala-Hypothalamic Circuit Involved in Food Reward. J Neurosci. 2019;39(5):888–99. Epub 2018/12/19. doi: 10.1523/JNEUROSCI.0250-18.2018. PubMed PMID: 30559151; PMCID: PMC6382977.

9. Fang Z, Spaeth AM, Ma N, Zhu S, Hu S, Goel N, Detre JA, Dinges DF, Rao H. Altered salience network connectivity predicts macronutrient intake after sleep deprivation. Sci Rep. 2015;5:8215. Epub 2015/02/04. doi: 10.1038/srep08215. PubMed PMID: 25645575; PMCID: PMC4314629.

10. Yang CL, Schnepp J, Tucker RM. Increased Hunger, Food Cravings, Food Reward, and Portion Size Selection after Sleep Curtailment in Women Without Obesity. Nutrients. 2019;11(3). Epub 2019/03/22. doi: 10.3390/nu11030663. PubMed PMID: 30893841; PMCID: PMC6470707.

11. Bayon V, Leger D, Gomez-Merino D, Vecchierini MF, Chennaoui M. Sleep debt and obesity. Ann Med. 2014;46(5):264–72. Epub 2014/07/12. doi: 10.3109/07853890.2014.931103. PubMed PMID: 25012962.

12. Strand LB, Laugsand LE, Wisloff U, Nes BM, Vatten L, Janszky I. Insomnia symptoms and cardiorespiratory fitness in healthy individuals: the Nord-Trondelag Health Study (HUNT). Sleep. 2013;36(1):99–108. Epub 2013/01/05. doi: 10.5665/sleep.2310. PubMed PMID: 23288976; PMCID: PMC3524509.

13. Chasens ER, Sereika SM, Weaver TE, Umlauf MG. Daytime sleepiness, exercise, and physical function in older adults. J Sleep Res. 2007;16(1):60–5. Epub 2007/02/21. doi: 10.1111/j.1365-2869.2007.00576.x. PubMed PMID: 17309764.

14. Holfeld B, Ruthig JC. A longitudinal examination of sleep quality and physical activity in older adults. J Appl Gerontol. 2014;33(7):791–807. Epub 2014/09/19. doi: 10.1177/0733464812455097. PubMed PMID: 25231754.

15. Haario P, Rahkonen O, Laaksonen M, Lahelma E, Lallukka T. Bidirectional associations between insomnia symptoms and unhealthy behaviours. J Sleep Res. 2013;22(1):89–95. Epub 2012/09/18. doi: 10.1111/j.1365-2869.2012.01043.x. PubMed PMID: 22978579.

16. Lambiase MJ, Gabriel KP, Kuller LH, Matthews KA. Temporal relationships between physical activity and sleep in older women. Med Sci Sports Exerc. 2013;45(12):2362–8. doi: 10.1249/MSS.0b013e31829e4cea. PubMed PMID: 23739529; PMCID: PMC3833970.

17. Baron KG, Reid KJ, Kern AS, Zee PC. Role of sleep timing in caloric intake and BMI. Obesity (Silver Spring). 2011;19(7):1374–81. Epub 2011/04/30. doi: 10.1038/oby.2011.100. PubMed PMID: 21527892.

18. Ogilvie RP, Patel SR. The epidemiology of sleep and obesity. Sleep Health. 2017;3(5):383–8. Epub 2017/09/20. doi: 10.1016/j.sleh.2017.07.013. PubMed PMID: 28923198; PMCID: PMC5714285.

19. Abeysena C, Jayawardana P. Stleep deprivation, physical activity and low income are risk factors for inadequate weight gain during pregnancy: a cohort study. J Obstet Gynaecol Res. 2011;37(7):734–40. Epub 2011/07/09. doi: 10.1111/j.1447-0756.2010.01421.x. PubMed PMID: 21736667.

20. Hill C, Lipsky LM, Betts GM, Siega-Riz AM, Nansel TR. A Prospective Study of the Relationship of Sleep Quality and Duration with Gestational Weight Gain and Fat Gain. J Womens Health (Larchmt). 2021;30(3):405–11. Epub 2020/09/19. doi: 10.1089/jwh.2020.8306. PubMed PMID: 32945728; PMCID: PMC7957376.

21. Merkx A, Ausems M, Bude L, de Vries R, Nieuwenhuijze MJ. Weight gain in healthy pregnant women in relation to pre-pregnancy BMI, diet and physical activity. Midwifery. 2015;31(7):693–701. Epub 2015/05/20. doi: 10.1016/j.midw.2015.04.008. PubMed PMID: 25981808.

22. Paulino DSM, Pinho-Pompeu M, Raikov F, Freitas-Jesus JV, Machado HC, Surita FG. The Role of Health-related Behaviors in Gestational Weight Gain among Women with Overweight and Obesity: A Cross-sectional Analysis. Rev Bras Ginecol Obstet. 2020;42(6):316–24. Epub 2020/07/01. doi: 10.1055/s-0040-1712132. PubMed PMID: 32604434.

23. Wallace ML, Buysse DJ, Redline S, Stone KL, Ensrud K, Leng Y, Ancoli-Israel S, Hall MH. Multidimensional Sleep and Mortality in Older Adults: A Machine-Learning Comparison With Other Risk Factors. J Gerontol A Biol Sci Med Sci. 2019;74(12):1903–9. Epub 2019/02/20. doi: 10.1093/gerona/glz044. PubMed PMID: 30778527; PMCID: PMC6853700.

24. Wallace ML, Lee S, Hall MH, Stone KL, Langsetmo L, Redline S, Schousboe JT, Ensrud K, LeBlanc ES, Buysse DJ, MrOs, Groups SOFR. Heightened sleep propensity: a novel and high-risk sleep health phenotype in older adults. Sleep Health. 2019;5(6):630–8. Epub 2019/11/05. doi: 10.1016/j.sleh.2019.08.001. PubMed PMID: 31678177; PMCID: PMC6993140.

25. Lehrer HM, Yao Z, Krafty RT, Evans MA, Buysse DJ, Kravitz HM, Matthews KA, Gold EB, Harlow SD, Samuelsson LB, Hall MH. Comparing polysomnography, actigraphy, and sleep diary in the home environment: The Study of Women’s Health Across the Nation (SWAN) Sleep Study. Sleep Adv. 2022;3(1):zpac001. Epub 20220219. doi: 10.1093/sleepadvances/zpac001. PubMed PMID: 35296109; PMCID: PMC8918428.

26. Haas DM, Ehrenthal DB, Koch MA, Catov JM, Barnes SE, Facco F, Parker CB, Mercer BM, Bairey-Merz CN, Silver RM, Wapner RJ, Simhan HN, Hoffman MK, Grobman WA, Greenland P, Wing DA, Saade GR, Parry S, Zee PC, Reddy UM, Pemberton VL, Burwen DR, National Heart L, Blood Institute nuMo MbHHSN. Pregnancy as a Window to Future Cardiovascular Health: Design and Implementation of the nuMoM2b Heart Health Study. Am J Epidemiol. 2016;183(6):519–30. Epub 2016/01/31. doi: 10.1093/aje/kwv309. PubMed PMID: 26825925; PMCID: PMC4782765.

27. Reid KJ, Facco FL, Grobman WA, Parker CB, Herbas M, Hunter S, Silver RM, Basner RC, Saade GR, Pien GW, Manchanda S, Louis JM, Nhan-Chang CL, Chung JH, Wing DA, Simhan HN, Haas DM, Iams J, Parry S, Zee PC. Sleep During Pregnancy: The nuMoM2b Pregnancy and Sleep Duration and Continuity Study. Sleep. 2017;40(5). Epub 2017/04/04. doi: 10.1093/sleep/zsx045. PubMed PMID: 28369543; PMCID: PMC6396817.

28. Buysse DJ. Sleep health: can we define it? Does it matter? Sleep. 2014;37(1):9–17. Epub 2014/01/29. doi: 10.5665/sleep.3298. PubMed PMID: 24470692; PMCID: PMC3902880.

29. Wallace ML, Yu L, Buysse DJ, Stone KL, Redline S, Smagula SF, Stefanick ML, Kritz-Silverstein D, Hall MH. Multidimensional sleep health domains in older men and women: an actigraphy factor analysis. Sleep. 2021;44(2). doi: 10.1093/sleep/zsaa181. PubMed PMID: 32918075; PMCID: PMC7879411.

30. Hirshkowitz M, Whiton K, Albert SM, Alessi C, Bruni O, DonCarlos L, Hazen N, Herman J, Adams Hillard PJ, Katz ES, Kheirandish-Gozal L, Neubauer DN, O’Donnell AE, Ohayon M, Peever J, Rawding R, Sachdeva RC, Setters B, Vitiello MV, Ware JC. National Sleep Foundation’s updated sleep duration recommendations: final report. Sleep Health. 2015;1(4):233–43. Epub 2015/12/01. doi: 10.1016/j.sleh.2015.10.004. PubMed PMID: 29073398.

31. Ohayon M, Wickwire EM, Hirshkowitz M, Albert SM, Avidan A, Daly FJ, Dauvilliers Y, Ferri R, Fung C, Gozal D, Hazen N, Krystal A, Lichstein K, Mallampalli M, Plazzi G, Rawding R, Scheer FA, Somers V, Vitiello MV. National Sleep Foundation’s sleep quality recommendations: first report. Sleep Health. 2017;3(1):6–19. Epub 2017/03/28. doi: 10.1016/j.sleh.2016.11.006. PubMed PMID: 28346153.

32. Bowman MA, Brindle RC, Joffe H, Kline CE, Buysse DJ, Appelhans BM, Kravitz HM, Matthews KA, Neal-Perry GS, Krafty RT, Hall MH. Multidimensional sleep health is not cross-sectionally or longitudinally associated with adiposity in the Study of Women’s Health Across the Nation (SWAN). Sleep Health. 2020;6(6):790–6. Epub 2020/07/19. doi: 10.1016/j.sleh.2020.04.014. PubMed PMID: 32680819.

33. Watson NF, Badr MS, Belenky G, Bliwise DL, Buxton OM, Buysse D, Dinges DF, Gangwisch J, Grandner MA, Kushida C, Malhotra RK, Martin JL, Patel SR, Quan SF, Tasali E. Recommended Amount of Sleep for a Healthy Adult: A Joint Consensus Statement of the American Academy of Sleep Medicine and Sleep Research Society. Sleep. 2015;38(6):843–4. Epub 2015/06/04. doi: 10.5665/sleep.4716. PubMed PMID: 26039963; PMCID: PMC4434546.

34. Cox JL, Holden JM, Sagovsky R. Detection of postnatal depression. Development of the 10-item Edinburgh Postnatal Depression Scale. The British journal of psychiatry: the journal of mental science. 1987;150:782–6. PubMed PMID: 3651732.

35. Kirkpatrick SI, Reedy J, Krebs-Smith SM, Pannucci TE, Subar AF, Wilson MM, Lerman JL, Tooze JA. Applications of the Healthy Eating Index for Surveillance, Epidemiology, and Intervention Research: Considerations and Caveats. J Acad Nutr Diet. 2018;118(9):1603–21. doi: 10.1016/j.jand.2018.05.020. PubMed PMID: 30146072; PMCID: PMC6730554.

36. Hutcheon JA, Platt RW, Abrams B, Himes KP, Simhan HN, Bodnar LM. A weight-gain-for-gestational-age z score chart for the assessment of maternal weight gain in pregnancy. Am J Clin Nutr. 2013;97(5):1062–7. Epub 20130306. doi: 10.3945/ajcn.112.051706. PubMed PMID: 23466397; PMCID: PMC3625243.

37. Goldstein RF, Abell SK, Ranasinha S, Misso M, Boyle JA, Black MH, Li N, Hu G, Corrado F, Rode L, Kim YJ, Haugen M, Song WO, Kim MH, Bogaerts A, Devlieger R, Chung JH, Teede HJ. Association of Gestational Weight Gain With Maternal and Infant Outcomes: A Systematic Review and Meta-analysis. JAMA. 2017;317(21):2207–25. Epub 2017/06/07. doi: 10.1001/jama.2017.3635. PubMed PMID: 28586887; PMCID: PMC5815056.

38. Chung J, Goodman M, Huang T, Bertisch S, Redline S. Multidimensional sleep health in a diverse, aging adult cohort: Concepts, advances, and implications for research and intervention. Sleep Health. 2021;7(6):699–707. Epub 2021/10/14. doi: 10.1016/j.sleh.2021.08.005. PubMed PMID: 34642124; PMCID: PMC8665047.

39. Brindle RC, Yu L, Buysse DJ, Hall MH. Empirical derivation of cutoff values for the sleep health metric and its relationship to cardiometabolic morbidity: results from the Midlife in the United States (MIDUS) study. Sleep. 2019;42(9). Epub 2019/05/15. doi: 10.1093/sleep/zsz116. PubMed PMID: 31083710; PMCID: PMC7458275.

40. McDonald SD, Yu ZM, van Blyderveen S, Schmidt L, Sword W, Vanstone M, Biringer A, McDonald H, Beyene J. Prediction of excess pregnancy weight gain using psychological, physical, and social predictors: A validated model in a prospective cohort study. PLoS One. 2020;15(6):e0233774. Epub 2020/06/03. doi: 10.1371/journal.pone.0233774. PubMed PMID: 32484813; PMCID: PMC7266315.

41. Hill B, Skouteris H, McCabe M, Milgrom J, Kent B, Herring SJ, Hartley-Clark L, Gale J. A conceptual model of psychosocial risk and protective factors for excessive gestational weight gain. Midwifery. 2013;29(2):110–4. Epub 2012/11/20. doi: 10.1016/j.midw.2011.12.001. PubMed PMID: 23159235.

42. Matthews J, Huberty J, Leiferman J, Buman M. Psychosocial predictors of gestational weight gain and the role of mindfulness. Midwifery. 2018;56:86–93. Epub 2017/11/03. doi: 10.1016/j.midw.2017.10.008. PubMed PMID: 29096284.

43. Ladyman C, Signal TL. Sleep Health in Pregnancy: A Scoping Review. Sleep Med Clin. 2018;13(3):307–33. doi: 10.1016/j.jsmc.2018.04.004. PubMed PMID: 30098750.

44. Carroll JE, Teti DM, Hall MH, Christian LM. Maternal Sleep in Pregnancy and Postpartum Part II: Biomechanisms and Intervention Strategies. Curr Psychiatry Rep. 2019;21(3):19. Epub 20190302. doi: 10.1007/s11920-019-1000-9. PubMed PMID: 30826895.

45. DeSantis AS, Dubowitz T, Ghosh-Dastidar B, Hunter GP, Buman M, Buysse DJ, Hale L, Troxel WM. A preliminary study of a composite sleep health score: associations with psychological distress, body mass index, and physical functioning in a low-income African American community. Sleep Health. 2019;5(5):514–20. Epub 2019/06/19. doi: 10.1016/j.sleh.2019.05.001. PubMed PMID: 31208939; PMCID: PMC6801051.

46. Balieiro LCT, Gontijo CA, Fahmy WM, Maia YCP, Crispim CA. Does sleep influence weight gain during pregnancy? A prospective study. Sleep Sci. 2019;12(3):156–64. Epub 2020/01/01. doi: 10.5935/1984-0063.20190087. PubMed PMID: 31890090; PMCID: PMC6932844.

47. Pauley AM, Hohman EE, Leonard KS, Guo P, McNitt KM, Rivera DE, Savage JS, Downs DS. Short Nighttime Sleep Duration and High Number of Nighttime Awakenings Explain Increases in Gestational Weight Gain and Decreases in Physical Activity but Not Energy Intake among Pregnant Women with Overweight/Obesity. Clocks Sleep. 2020;2(4):487–501. Epub 2020/11/19. doi: 10.3390/clockssleep2040036. PubMed PMID: 33202691; PMCID: PMC7711788.

